# Routine SARS-CoV-2 wastewater surveillance results in Turkey to follow Covid-19 outbreak

**DOI:** 10.1101/2020.12.21.20248586

**Authors:** Bilge Alpaslan Kocamemi, Halil Kurt, Ahmet Sait, Hamza Kadi, Fahriye Sarac, Ismail Aydın, Ahmet Mete Saatci, Bekir Pakdemirli

**Affiliations:** Marmara University, Department of Environmental Engineering, Istanbul, Turkey; Saglik Bilimleri University, Hamidiye International School of Medicine, Department of Medical Biology, Istanbul, Turkey; Ministry of Agriculture and Forestry, Republic of Turkey, Veterinary Control Central Research Institute, Pendik, Istanbul, Turkey; Ministry of Agriculture and Forestry, Republic of Turkey, Veterinary Control Central Research Institute, Samsun, Turkey; Turkish Water Institute, Istanbul, Turkey; Ministry of Agriculture and Forestry, Republic of Turkey, Ankara, Turkey

**Author notes:** **Bilge Alpaslan Kocamemi:** Methodology, Data Curation, Formal analysis, Writing - Original Draft, Preparation, Writing - Review & Editing, Visualization, Supervision, Project administration. **Halil Kurt:** Validation, Verification, Data Curation, Writing - Review & Editing. **Ahmet Sait, Hamza Kadi:** Investigation, **Fahriye Sarac, Ismail Aydin:** Validation, **Ahmet Mete SaatçI** : Conceptualization, Writing - Review & Editing, Supervision, Project administration. **Bekir Pakdemirli:** Resources, Funding acquisition.

**Keywords:** Colored-scale map, Covid-19, nationwide surveillance, SARS-CoV-2, viral load

## Abstract

A global pandemic of Coronavirus Disease 2019 (Covid-19) caused by severe acute respiratory syndrome coronavirus 2 (SAR-CoV-2) declared by WHO in March 2019 is still ongoing. As of 13^th^ of December 2020, 70 million people were infected by SARS-CoV-2 and 1.5 million people lost their lives globally (WHO, 2020). Since March 2019, diagnosis of Covid-19 cases has been done through PCR test of samples from nasopharyngeal and throat swabs. However, in March 2019, it was reported that the faeces [1] and urine [2] of all infected people contain SARS-CoV-2. Later, numerous researchers [3-7] detected SARS-CoV-2 in faeces of both symptomatic and asymptomatic patients. Moreover, some studies [1,4,8-12] suggested the possibility of extended duration of viral shedding in faeces after the patients’ respiratory samples tested negative. In this respect, SARS-CoV-2 wastewater-based epidemiology (WBE), i.e., wastewater surveillance, aiming to estimate the distribution of asymptomatic and symptomatic individuals in a specific region has received worldwide attention. Various research groups worldwide [1, 13-54] have started SARS-CoV-2 detection in wastewater since WBE provides tracking whole population by testing a small number of wastewater samples in a specific region and can predict SARS-CoV-2 RNA in human faeces a few days to a week before onset of symptoms. This makes WBE quite economic tool for continual tracking of decreasing or increasing trend of the Covid-19 in a particular region. However, up to date, almost all of the WBE studies have been performed with samples from a few treatment plants. There was no reported nationwide wastewater surveillance study that has been integrated into a national Covid-19 management strategy by decision makers. Nationwide, SARS-CoV-2 surveillance studies have great potential to reflect the actual distribution of Covid-19 cases in a community by accounting not only symptomatic patients tested but also asymptomatic patients having no or mild symptoms and not been tested. As opposed to clinical surveillance studies, wastewater-based surveillance studies will reflect the number of cases in a community by testing one sample from a treatment plant serving this community instead of performing individual swab tests.

Turkey, which is among the few countries that started wastewater based surveillance studies at the early stages of pandemic is a leading country, performing a nationwide surveillance study. The distribution of Covid-19 cases throughout the country via SARS-CoV-2 measurements in influent, effluent and sludge samples of wastewater treatment plants (WWTPs) located in 81 cities through May 2020-June 2020 was conducted [36, 51, 52]. In June 2020, nationwide routine sampling through 22 regional identified cities has been started. However, from June to September 2020 all samples were detected negative due to problems with RT-pCR primer targeting RdRp gene of SARS-CoV-2 genome. Since September 2020, routine sampling from 22 cities of Turkey with 2 weeks sampling period (weekly for mega city Istanbul) has been continued and regional Covid-19 distributions have been reported as viral loads on color-scale maps. To the best our knowledge, this is the first routine nationwide surveillance study indicating Covid-19 distribution regularly using color-scale presentation on a map.

## 2. Value of the Data

- A nationwide SARS-CoV-2 surveillance study has been routinely performed first time worldwide
- The measured virus concentrations have been normalized with actual flowrate of WWTP during sampling.
- Covid-19 distribution in a whole country has been demonstrated on a color-scaled map first time worldwide.

## 3. Methodology

### 3.1. Sampling

Routine nationwide wastewater SARS-COV-2 surveillance has been performed in 22 cities of Turkey (Fig. 1). These cities were selected as pilot cities based on population and Covid-19 potentials in regions identified by Turkey Ministry of Health. In these pilot cities, samples were collected at two-week intervals from the influents of the treatment plants serving the maximum population in that city. For mega-city Istanbul, weekly influent samples were collected from two major treatment plants (Fig. 2): Ambarlı WWTP located in the European side of Istanbul and serving about 1.4 million people, Pasakoy WWTP located in the Asian side of Istanbul and serving about 1 million people.

**Fig 1.**
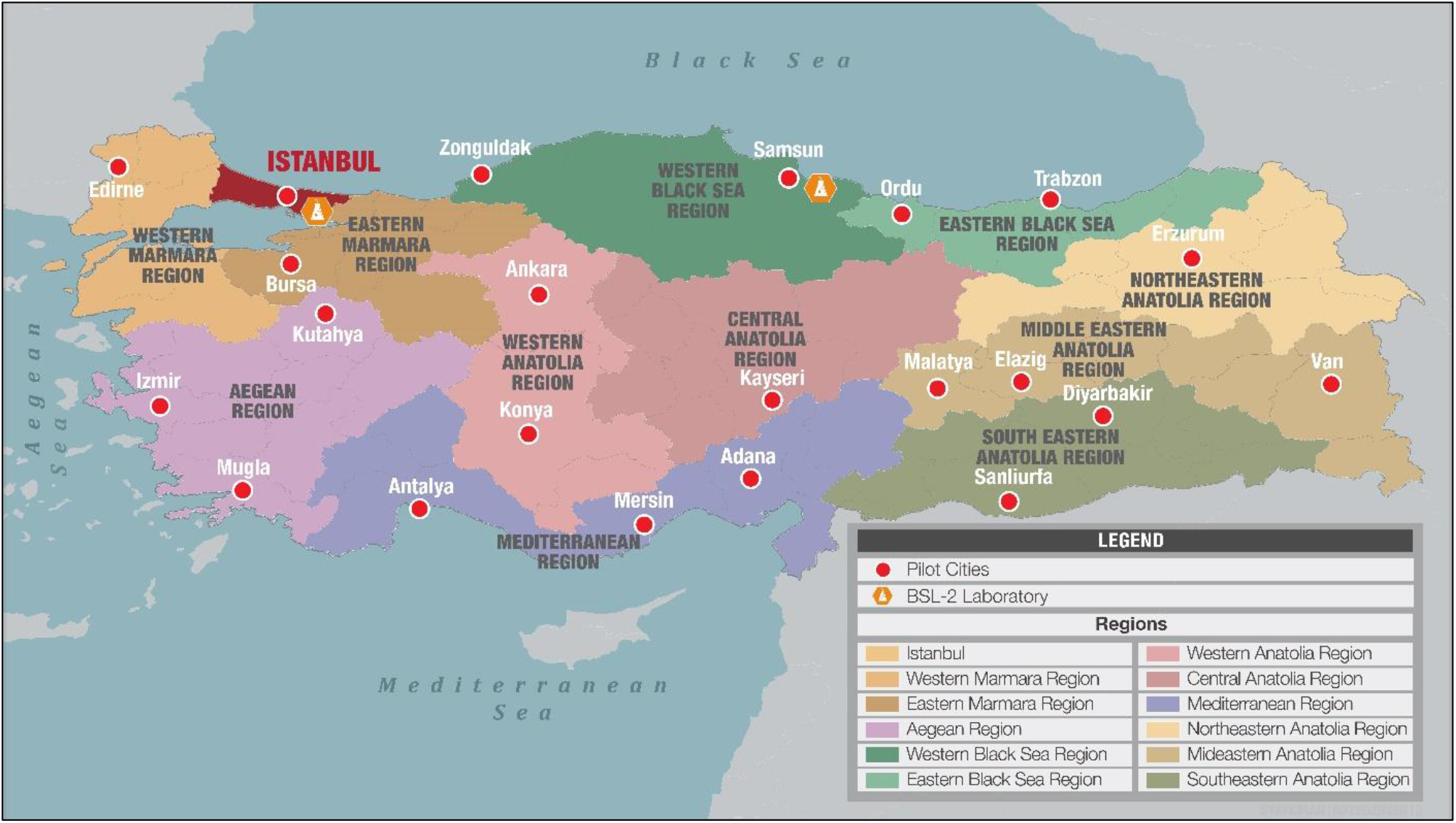
Routine **s**ampling locations in Turkey for nationwide SARS-CoV-2 surveillance study in wastewater.

**Fig 2.**
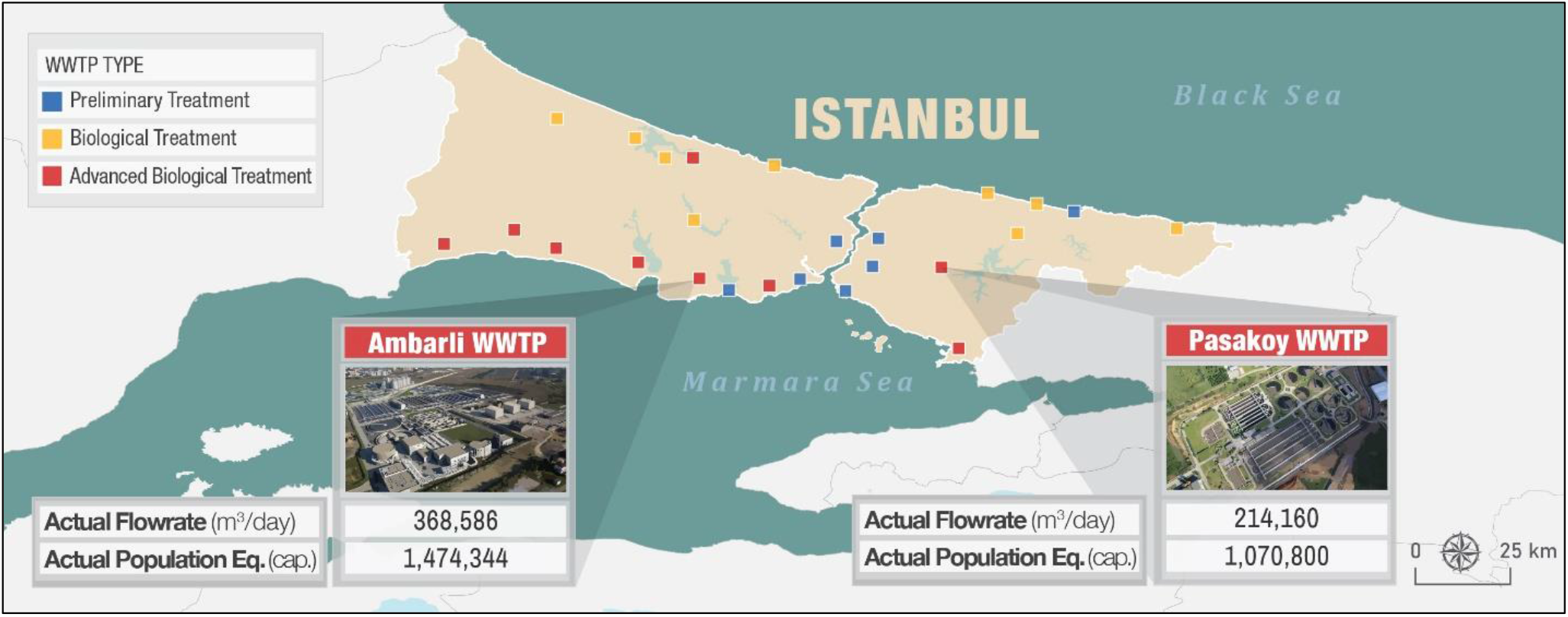
Routine **s**ampling locations in mega city Istanbul for nationwide SARS-CoV-2 surveillance study in Turkey.

### 3.2. SARS-CoV-2 concentration

Samples were shaken at 4°C at 100 rpm for 30 min in order to transfer attached viruses to the aqueous phase. Microorganisms and large particles were removed from the samples by centrifugation at 7471G for 30 minutes at 4 degrees celcius. 250 ml of supernatant was filtered through 0.45 μm and 0.2 μm to remove remaining particles and cell debris. Filtrate was mixed thoroughly with PEG 8000 (10% w/v) by shaking for 1 minute. The mixture was incubated at 4°C at 100 rpm for overnight. Following to the incubation, the mixture was divided in six 50 ml falcon tubes. Viruses were precipitated by centrifugation at 7471G for 120 minutes at 4 degrees celcius. Supernatant was removed carefully without disturbing the pellets. Pellets of each falcon tubes were re-suspended with 200 μl RNA free water. 1 ml of virus concentrate was used for total RNA extraction and remaining concentrate stored at -80°C. Total viral RNA was extracted either with Roche MagNA pure LC total nucleic acid isolation kit using Roche MagNA pure LC system in accordance with the manufacturer’s protocols (Istanbul Pendik Lab) or manual RNA extraction method (Samsun Lab). RNA was determined both qualitatively and quantitatively by Thermo NanoDrop 2000c (Penzberg, Germany). Due to high community risk of using SARS-CoV-2 virus, 300 µl of 10^5^ copy/ml surrogate avian coronavirus of Infectious Bronchitis Virus were added our samples in order to evaluate the virus recovery efficiency of PEG 8000 adsorption method. Based on RT-QPCR results, 1-1.5 log virus titer loss were observed after PEG 8000 adsorption and RNA isolation.

### 3.3. Quantitative reverse transcription PCR (RT-qPCR)

Primers and taqman probe sets targeting SARS-CoV-2 N gene [53] were used to detect and quantify SARS-CoV-2 virus. Serial dilutions of synthetic SARS-CoV-2 N gene were used as a standard for absolute quantification. RT-qPCR analysis was performed in Realtime ready RNA virus Master (Roche Diaonostics, Mannhaim, Germany) contained 0.8 nM of forward primer and reverse primer, 0.25 nM probe and 5 μL of template RNA. The RT-qPCR assays were performed at 50 °C for 6 min, 53 °C for 4 min, 58 °C for 4 min for reverse transcription, followed by 95 °C for 2 min and then 45 cycles of 95 °C for 10 s, 55 °C for 10 s and 72 °C for 1 s for data collection using a Roche LightCycle 2.0 thermal cycler (Roche Diaonostics, Mannhaim, Germany).

## 4. Data Description

### 4.1. Viral Load Calculation and Color-Scale Presentation on Map

The SARS-CoV-2 RT-qPCR measurements repeated three times for each sample and results (virus titer/l) were normalized based on the actual flowrates measured at WWTPs in order to present daily viral loads. By considering the minimum and maximum detection limits of RT-qPCR measurements together with minimum and maximum actual flowrates in the pilot cities, the minimum (1E+8 virus titer/day) and the maximum (1E+14 virus titer/day) viral load values, which are possible to observe, were identified. The identified range was then sub-classified under 4 groups on color-scale (Figure 3) as High Case group, Moderate Case group, Low Case group, Very Low Case/ No Case group.

**Fig 3.**
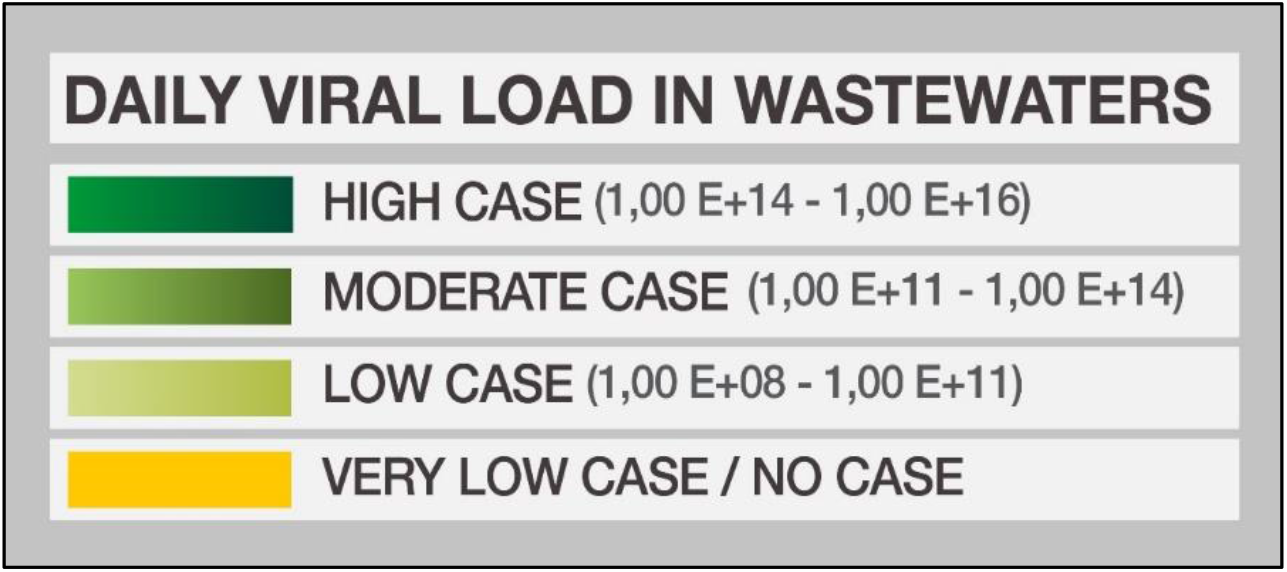
Viral load presentation based on Covid -19 distributions on color-scale.

### 4.2. Distribution of Covid-19 Cases in Turkey Based on Observed Viral Loads in Wastewater

The observed viral loads in 21 pilot cities of Turkey are shown color-scale maps in Figure 4. At the beginning of pandemic between May 2020 and June 2020, the Covid-19 cases were mostly in the western part of Turkey, especially in Istanbul, Bursa and Konya in the Middle Anatolian region of Turkey. At that time, Covid-19 cases were low in pilot cities located in the Aegean, Mediterranean and Black Sea regions. Cases were significantly low or absent in the Eastern and Southeastern regions. During routine sampling period, from 15^th^ of October to 15^th^ of November 2020, Covid-19 cases started to spread in low numbers over Middle Anatolian, Mediterranean, Eastern and Southeastern areas. In the period of 15^th^ of November to 15^th^ of December 2020, the Covid-19 cases were distributed over the whole country and reached to moderate levels in the Middle Anatolian, Eastern and Southeastern regions where no or very low cases were observed before November.

**Fig 4.**
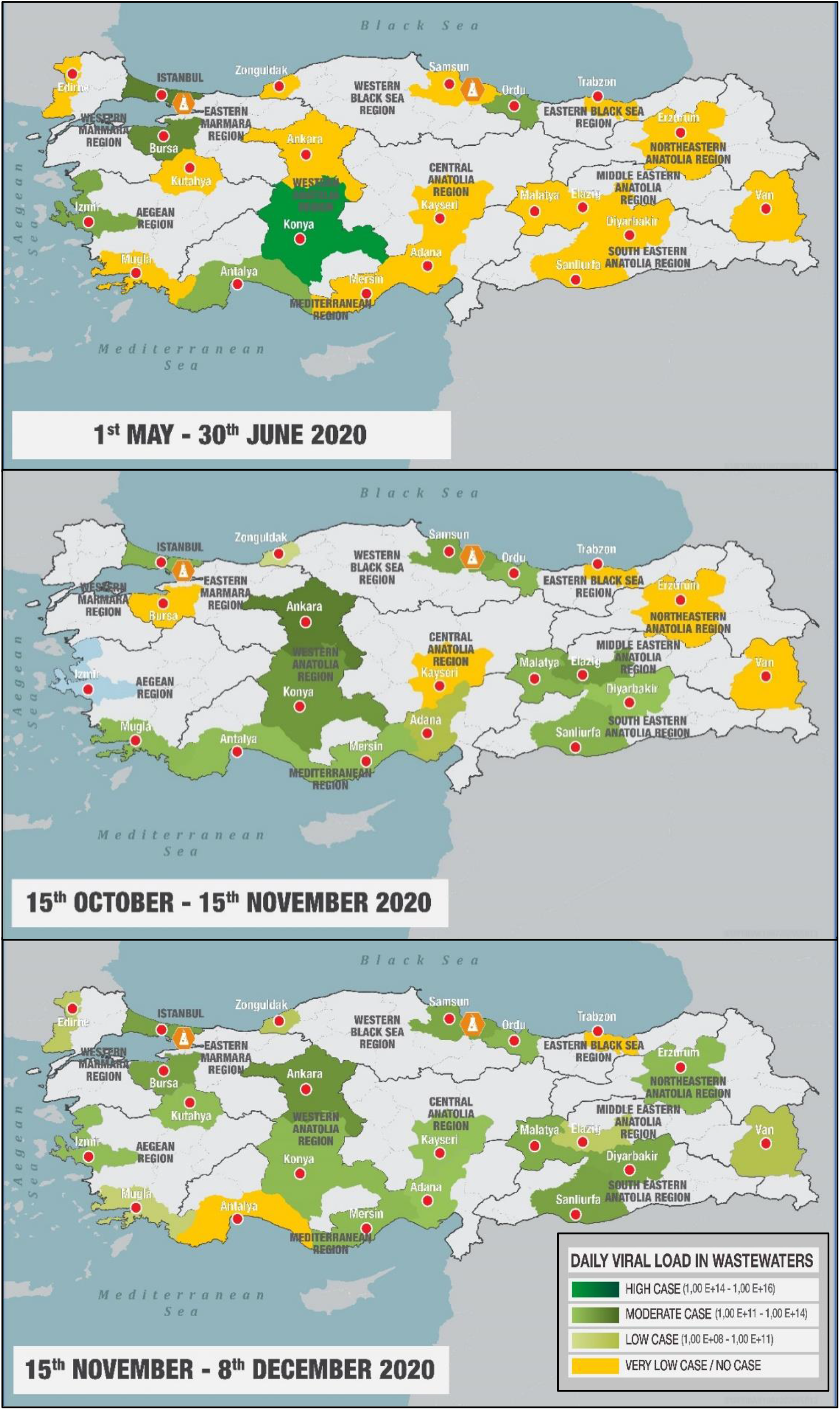
Change of SARS-CoV-2 viral loads throughout Turkey with time.

### 4.3 Distribution of Covid-19 Cases in mega-city Istanbul Based on Observed Viral Loads in Wastewater

Since the start of pandemic in Turkey (April 2020), Covid-19 cases have been mostly prevailing in the mega-city of Istanbul with 15 million inhabitants. Figure 5 presents the observed viral loads in Istanbul wastewater since the start of the pandemic. Based on viral loads, Covid-19 cases were very high in April and in May 2020. From June to August 2020, SARS-CoV-2 could not be detected in wastewater due to RT-qPCR primer problem explained under Section 1. Starting from August 2020, viral load changes revealed that Covid-19 cases started to increase in October 2020 and reached to a peak in November 2020 resulting in government lock-downs after 9:00 PM and weekends. The positive effect of lockdowns was apparent on viral load map of 1^st^ week of December 2020 which shows the successful applicability of wastewater-based Covid-19 surveillance.

**Fig 5.**
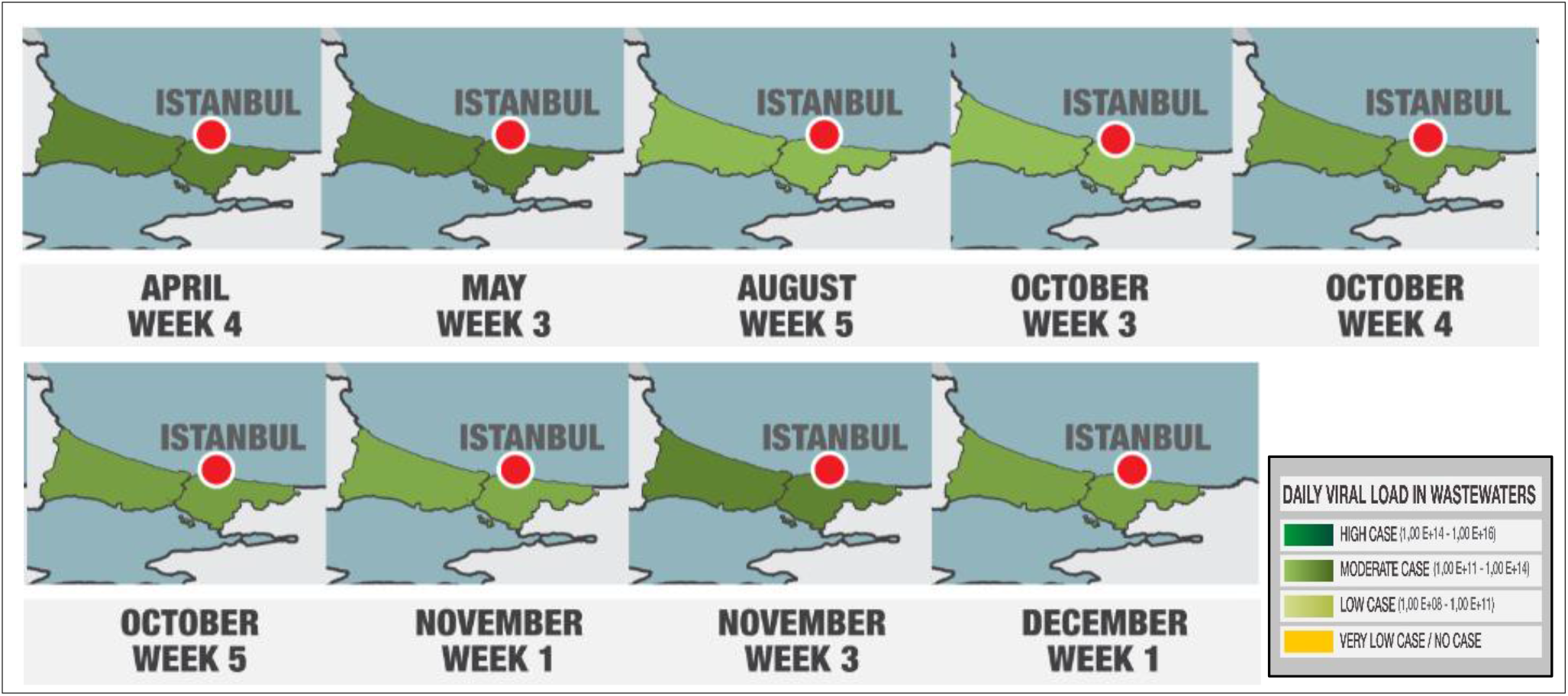
Change of SARS-CoV-2 viral load distributions in Istanbul mega city.

### 4.4. Correlation of Observed Viral Loads in Wastewater with Case Numbers

The routine nationwide wastewater surveillance study results of Turkey were further evaluated based on the reported case numbers (Fig. 6). Since the number of cases have been reported regionally, the case numbers shown in Fig. 6 belong to the region of the city that is located. However, while the case number trends vary with viral load changes, it is apparent that viral load trends do not correspond with the change in case numbers. Although, measured viral loads were high in October and did not change significantly in November, the number of reported cases demonstrated an increase from October to November. This observation may indicate the presence of significant numbers of asymptomatic patients in October and may also show the presence of SARS-CoV-2 RNA in human feaces a few days to a week before onset of symptoms.

**Fig 5.**
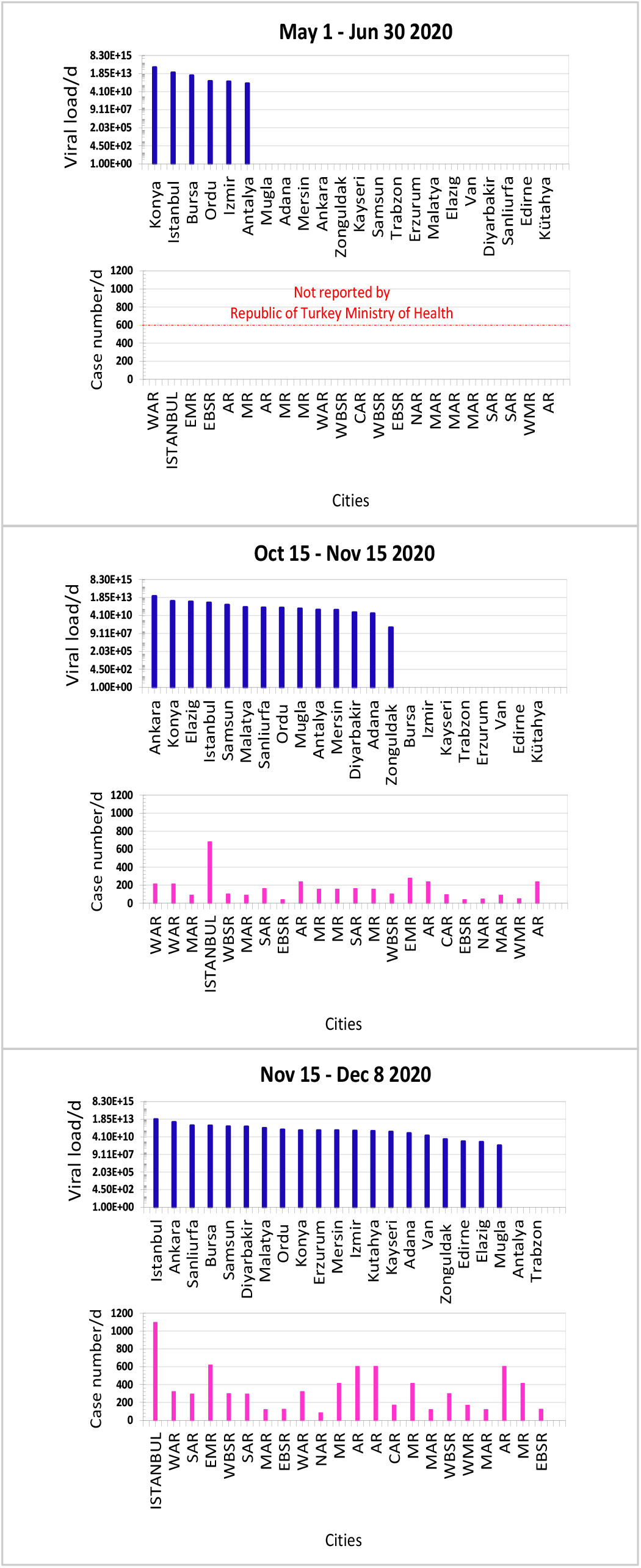
City-based SARS-CoV-2 viral load distributions and regional reported case numbers [54]. WMR: Western Marmara Region, EMR: Eastern Marmara Region, WAR: Western Anatolia Region, CAR: Central Anatolia Region; AR: Aegean Region, WBSR: Western Black Sea Region, EBSR: Eastern Black Sea Region, MR: Mediterranean Region, MAR: Mideastern Anatolia Region, NAR: Northeastern Anatolia Region, SAR: Southeastern Anatolia Region

## Data Availability

All data is available

## Ethics Statement

The work did not involve any human subject and animal experiments.

### Acknowledgments

This work was financed by Republic of Turkey, Ministry of Agriculture and Forestry.

The authors wish to acknowledge the Turkish Water Institute (SUEN) for the coordination and execution of this study. We wish to express our appreciation to the Ministry’s Istanbul Pendik and Samsun Veterinary Control Central Research Institute for their hard work and rapid analysis of the wastewater samples. We thank to DSI (State Hydraulic Works) for their logistic support for intercity sample transportation. We also thank to all municipalities for their cooperation and efforts to collect and preserve the sewage samples rapidly. Our special thanks to Env. Eng. Salim Yaykıran from SUEN for regional maps and Env.Eng. Sumeyye Celik from Marmara University her contribution to gather and collate up to date information about the worldwide studies on SARS-CoV-2 in wastewater.

